# Relation of Self-Reported Race and Genetic Ancestry to Hypertension Prevalence Among Hispanics/Latinos: The Hispanic Community Health Study/Study of Latinos

**DOI:** 10.64898/2026.09.01.26361995

**Authors:** Raul A Montanez-Valverde, Vivian Kim, Priscilla A Duran Luciano, Yawen Yuan, Tamar Sofer, Robert Kaplan, Linda C Gallo, Gregory A Talavera, Krista M Perreira, Martha Daviglus, Sylvia E Rosas, Maria M Llabre, Tali Elfassy, Xihao Li, Carmen R Isasi, Carlos J Rodriguez

**Author notes:** **Corresponding author:** Raul A. Montanez-Valverde, MD, MS, Address: Albert Einstein College of Medicine. Jack and Pearl Resnick Campus. 1300 Morris Park Avenue. Block 509. Bronx, NY 10461.

## Abstract

**Background:** The imprecision of current metrics to capture the complex genetic admixture and racial identity among Hispanic/Latino individuals in the United States (US) is a concern. We examined the relationship of self-reported race and genetic ancestry with hypertension (HTN) among Hispanics/Latinos.

**Methods:** Cross-sectional study of the Hispanic Community Health Study/Study of Latinos (HCHS/SOL), including 10,586 Hispanic/Latino unrelated adults. Genetic ancestry: West African (AA), Amerindian (AI), and European (EA). Self-reported race: White, Black, Native American, or Multiple/Missing (*More than one race* or *Unknown/Not reported/Refused*). HTN: systolic (SBP) ≥130 mmHg, diastolic blood pressure (DBP) ≥80 mmHg, and/or use of HTN medications. Age- and sex adjusted models were used.

**Results:** Self-reported race was White (38·6%), Black (3·6%), Native American (4·1%), and Multiple/Missing (53·7%), with *Unknown/Not reported/Refused* representing 32·7%. Black and White Hispanics/Latinos had the greatest AA (55·7%) and EA (69·3%) ancestries, respectively. Each 10% AA increase was associated with OR 1·15, SBP beta +0.9 mmHg, and DBP beta +0.7 mmHg. Conversely, each 10% AI increase was associated with OR 0·83, SBP beta -0·4 mmHg, and DBP beta -0·6 mmHg. HTN prevalence was highest among those with Black race or in the highest AA quantile (45·6% and 48·0%, respectively), and lowest among those with Native American race or in the highest AI quantile (37·6% and 26·7%, respectively).

**Conclusion:** One-third of Hispanics/Latinos did not self-report race. Black or White self-reporting race did somewhat relate to AA or EA ancestry, respectively. HTN profiles were related to self-reported race and genetic ancestry in this admixed population.

## INTRODUCTION

The histories of Latin American colonization and migration resulted in unique racial and genetic admixture among the Hispanic/Latino population.^1^ While Hispanics/Latinos are often considered as a single ethnic group in research and governmental matters, they actually have significant racial heterogeneity.^2^ Among Hispanic/Latino adults living in the United States (US), race is often either not recognized or conflated with ethnicity.^1,3^ Further, some Hispanic/Latino adults may perceive the US concept of race as incompatible with their ethnic identity.^4^ Thus, historically, many do not self-identify with any particular race^1,5,6^, choose “Unknown/Other”, or do not answer when asked to self-select a race.^4–6^ This is not surprising, since race is socially constructed, and the social context of race in Latin America differs from that in the US.

Self-reported race, among non-Hispanics in the US, is associated with differential healthcare outcomes and social determinants of health (SDoH).^7^ Hypertension (HTN) prevalence is higher in non-Hispanic Black (NHB) than non-Hispanic White (NHW) persons.^8^ Among Hispanic/Latino people, HTN prevalence is higher in those of Caribbean background (Dominican, Cuban, and Puerto Rican),^3,9^ a group with higher African ancestry compared with Mainland heritage groups.^10^ However, HTN traits among Hispanics/Latinos who self-identify race as Black or Native American and those with African or Amerindian ancestry remain poorly characterized.

We examined data from the Hispanic Community Health Study/Study of Latinos (HCHS/SOL), the largest comprehensive epidemiologic cohort study focusing on Hispanic/Latino adults in the US.^11^ Our aims were to 1) determine the correspondence between self-reported race and genetic ancestry among Hispanics/Latinos; 2) examine the association of genetic ancestry and self-reported race with HTN prevalence among Hispanics/Latinos; 3) determine the aforementioned relationships across disaggregated Hispanic/Latino background/heritage groups.

## METHODS

HCHS/SOL is a multicenter, population-based, prospective cohort study that enrolled 16,415 Hispanic/Latino adults at baseline from randomly selected households in four US metropolitan cities (Chicago, Illinois; Miami, Florida; Bronx, New York; and San Diego, California) between 2008 and 2011. The specific study design and aims of HCHS/SOL are reported elsewhere^12^, and all questionnaires were administered in the preferred language (English or Spanish). HCHS/SOL included adults who answered yes to the question: “*Do you consider yourself to be Hispanic/Latino?”*

### Participants

Two levels of analysis with different sample sizes were performed. First, for the overall analysis, of the 16,415 participants in HCHS/SOL visit 1, those who self-reported race as *Asian, Native Hawaiian, or Other Pacific Islander* (n=99) were excluded due to their small representation, yielding a final analytic sample of 16,316 participants. Second, for the genetic ancestry-stratified analysis, from the 12,803 participants who provided consent for genetic studies at baseline, 10,642 were mutually unrelated, yielding a final analytic sample of 10,586 participants after some missingness.

### Hispanic/Latino background (heritage) groups

Obtained from the question: “*Which of the following best describes your Hispanic/Latino heritage: Dominican, Cuban, Puerto Rican, Central American, Mexican, South American, More than one heritage, and Other*.” We characterized *Dominican, Cuban,* and *Puerto Rican* as Caribbean background groups; and *Central American, Mexican,* and *South American* groups as Hispanic/Latinos of Mainland origin because of their shared anthropological history. *More than one heritage* was represented separately. We excluded those who selected “Other” (n=101) or “Refused to answer” (n=8) due to small representation and those with missing values (n=79).

### Self-reported race

Obtained based on the question: “*In addition to being of Hispanic/Latino heritage, which of the following categories would you use to describe yourself? (Mark only one): American Indian or Alaskan Native, Asian, Native Hawaiian or Other Pacific Islander, Black or African American, White, More than one race, Unknown or Not reported, and Refused*.” *American Indian or Alaskan Native* are considered Native American. For our study, we defined a Multiple/Missing self-reported race category as a composite of reporting: 1) *More than one race* or 2) *Unknown, Not reported, or Refused*. *Asian, Native Hawaiian, or Other Pacific Islander* (n=99) were excluded because of their small representation.

### Continental genetic ancestry

DNA extracted from blood samples was genotyped in HCHS/SOL using the Illumina Omni 2·5M array.^10^ West African (AA), Amerindian (AI), and European (EA) continental genetic ancestry proportions were estimated using ADMIXTURE.^13^ East Asian ancestry was not reported due to the low proportion.

### Outcomes

Blood pressure (BP) was reported as the average of three seated measurements obtained after a 5-minute rest. HTN was defined by: systolic blood pressure (SBP) ≥130 mmHg, diastolic blood pressure (DBP) ≥80 mmHg and/or self-reported use of antihypertensive medications.

### Statistical Analysis

Complex survey procedures and sampling weights were performed to account for the complex sampling design. Unweighted absolute counts characterized the study sample, while weighted percentages and means characterized the target Hispanic/Latino population. Mean, standard error (SE), and 95% confidence intervals (CI) were calculated for continuous variables, whereas percentages were reported for categorical variables. We presented the distribution of Hispanic/Latino background groups across self-reported races, as well as the distribution of self-reported race among Hispanic/Latino background groups. We calculated the χ-square statistic for multiple HTN proportions across self-reported races and performed an analysis of variance on the means of genetic ancestry proportions across races. For stratification analysis by genetic ancestry, we categorized participants as above or below the 80^th^ percentile and performed χ-square tests to compare HTN prevalence, and Student t-tests for two-group comparisons of SBP and DBP levels across these groups. Crude and age-/sex-adjusted logistic and linear regression analyses were conducted to assess the association between genetic ancestry proportion and HTN risk, as well as SBP and DBP values.

We performed several secondary analyses: 1) comparing subcomponents *More than one* race, and *Unknown/Not reported/Refused*; 2) utilizing the Seventh Report of the Joint National Committee (JNC 7) HTN definition: SBP ≥140 mmHg, DBP ≥90 mmHg; and 3) evaluating other genetic ancestry thresholds (50^th^ and 75^th^). All statistical analyses were performed using SAS survey procedures (SAS version 9.4, SAS Institute, Cary, NC). Statistical significance was defined as a two-tailed p-value < 0·05. Figures were created using GraphPad Prism version 8.0.0 for Windows and Biorender.com.

## RESULTS

The study population was 50·0% female, with a mean age of 42·9 (±0·3) years, a mean SBP of 121·0 (±0·3) mmHg, a mean DBP of 72·7 (±0·2) mmHg, and an HTN prevalence of 38·5%. **(Table 2 and eTable 1)**

### Self-reported race differences across Hispanic/Latino background groups

More than half of Hispanic/Latino persons (53·7%) overall classified as Multiple/Missing race. This was the case across background groups, except among Cuban heritage individuals, where 76·8% self-identified as White. **(Figure 1)** Within the Multiple/Missing race category, 34·9% reported *More than one race* and 65·3% reported *Unknown/Not reported/Refused*, corresponding to 17·5% and 32·7% of the overall study population, respectively. **(eTable 2)** White was the second most commonly self-reported race among individuals of South American, Central American, Mexican, and Puerto Rican descent, whereas Black was the second most common self-reported race among those of Dominican descent. **(Figure 1)** Self-reported race Black persons were primarily of Cuban or Dominican descent; self-reported White individuals were mainly Cuban or Mexican descent; and self-reported Native American people were predominantly of Mexican and Puerto Rican descent. **(Figure 1)** Persons of Mexican descent comprised the largest share of the Multiple/Missing category (43·3%) and represented over half of the *Unknown/Not reported/Refused* subcomponent. **(eTable 2)** eTable 4 shows a detailed breakdown of self-reported race and genetic ancestry by Hispanic/Latino background group.

**Figure 1.**
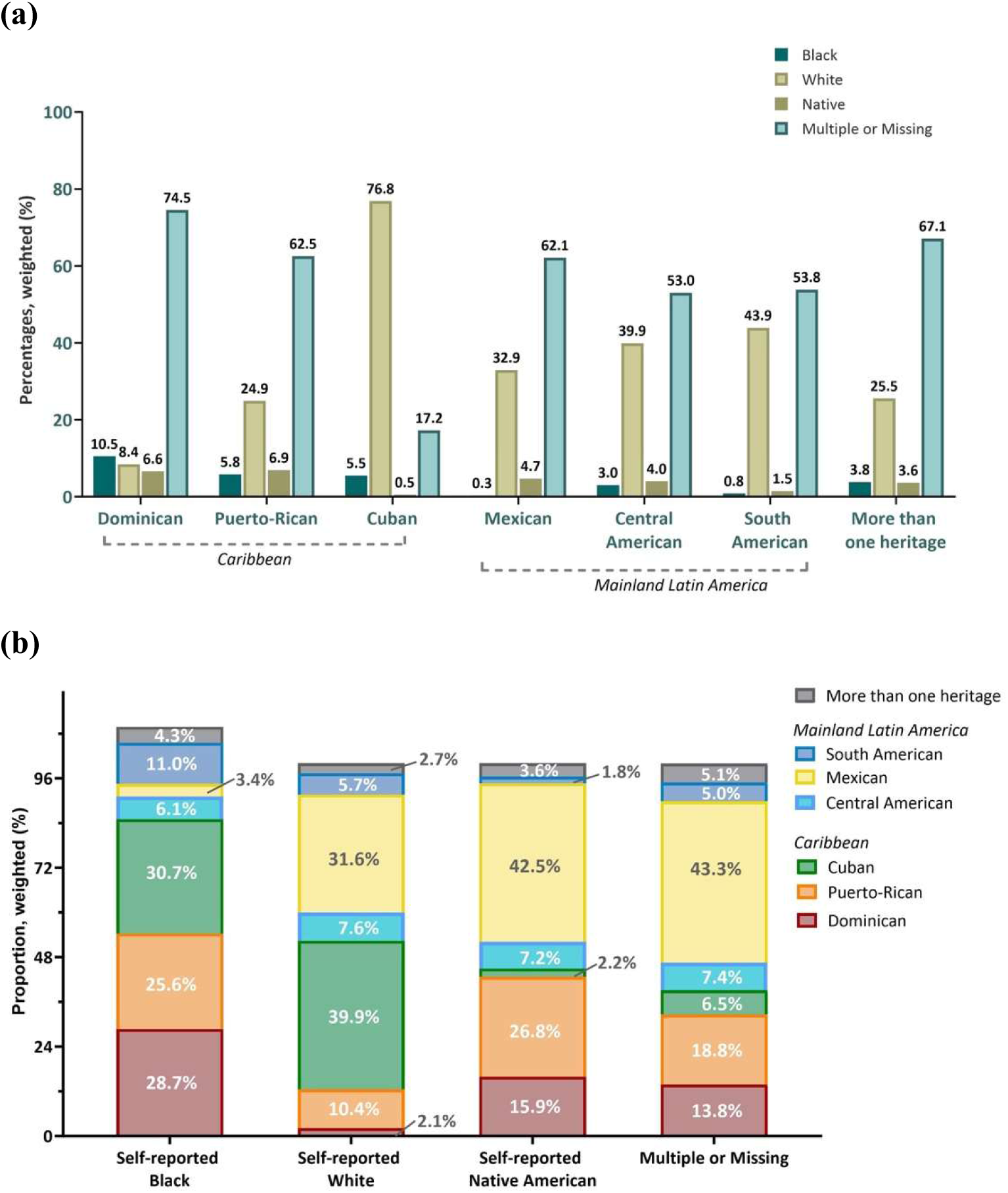
Proportions of Self-Reported Race Across Hispanic/Latino Background Group (a) and Proportions of Background Group Across Self-Reported Race (b). N=16,316 (N is unweighted, % are weighted). See Methods for additional information.

### Ancestry-predominance among self-reported race Black and White people

Overall, Hispanic/Latino persons who self-reported race as Black and White had the greatest proportions of AA and EA ancestry, respectively. **(Table 1)** Among Hispanics self-reporting Black race, 91·7% were in the highest AA quintile, whereas 41·6% of those who self-reported race as White were in the highest EA quintile. **(eTable 3).** This pattern remained consistent across Caribbean and Mainland background groups, but not for Hispanics/Latinos of Mexican descent. **(Figure 2, eTable 4)**

**Figure 2.**
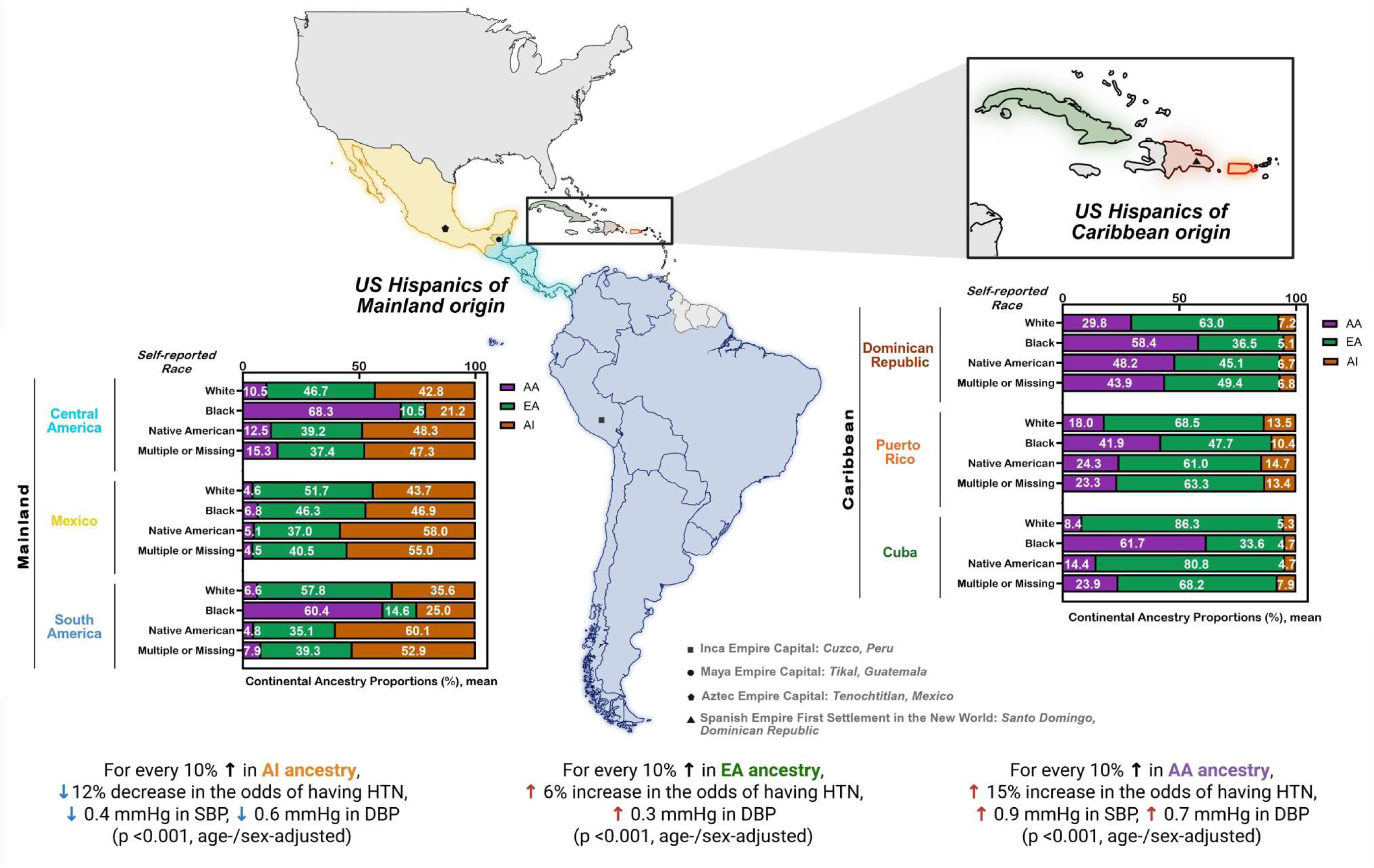
Genetic Ancestry and Self-Reported Race in US Hispanic-Latino Persons. AA: African ancestry, EA: European ancestry, AI: Amerindian ancestry, SBP: systolic blood pressure, DBP: diastolic blood pressure, HTN: hypertension. N=10,586 (N is unweighted, proportions and means are weighted). See Methods for additional information.

**Table 1.** Percent Genetic Ancestry Across Self-Reported Race Categories.

| Self-Reported Race | Overall,<br>N=16,316 |  | Genetic Ancestry proportion, N=10,586 |  |  |  |  |  |  |
| --- | --- | --- | --- | --- | --- | --- | --- | --- | --- |
|  | N | % | N | AA |  | EA |  | AI |  |
|  |  |  |  | Mean | SE | Mean | SE | Mean | SE |
| White | 6,021 | 38·6 | 4,114 | 8·8% | 0·2% | 69·3% | 0·8% | 22·0% | 0·8% |
| Black | 507 | 3·6 | 361 | 55·7% | 1·4% | 36·4% | 1·2% | 7·9% | 0·5% |
| Native American | 622 | 4·1 | 410 | 18·0% | 1·5% | 46·0% | 1·8% | 36·0% | 2·8% |
| Multiple/Missing | 9,166 | 53·7 | 5,701 | 17·2% | 0·6% | 48·7% | 0·5% | 34·1% | 0·7% |
|  | 16,316 |  | 10,586 | <0·001* |  | <0·001* |  | <0·001* |  |

N are unweighted, % (percentages) and means are weighted. SE: standard error; C.I.: confidence interval; AA: African ancestry; EA: European ancestry; AI: Amerindian ancestry; \*ANOVA p-value for means of proportions of continental genetic ancestry across self-reported races.

### Ancestry-mix among self-reported race as Native American or Multiple/Missing

Hispanic/Latino persons who self-reported race as Native American or Multiple/Missing had predominantly EA and AI ancestry **(Table 1)**, with higher mean AI proportions than any other self-reported race category. This pattern was consistent among Mainland Hispanics/Latinos **(Figure 2, eTable 4)**; however, among Caribbean Hispanics/Latinos, those who self-identified as Native American or Multiple/Missing had predominant EA and AA ancestry.

### Relation of Self-reported race and Genetic Ancestry to HTN prevalence and BP levels

HTN prevalence and BP levels were highest among Hispanic/Latino persons who self-reported race as Black and lowest among those who self-reported race as Native American or Multiple/Missing (both p-value <0·001). **(Table 2)** Similar patterns were observed using the JNC 7 HTN definition (*data not shown*). People in the highest AI quintile had lower HTN prevalence and BP levels than those with the lowest AI quintiles (p-value <0·001). In contrast, the highest HTN prevalence and BP levels were observed among individuals in the highest EA and AA quintiles compared with those in the respective lower quintiles (p < 0·001). **(Table 2)** Sensitivity analyses using the 50^th^ and 75^th^ genetic ancestry percentile thresholds yielded similar conclusions. **(eTable 5)** After adjusting for age and sex, each 10% increase in AA ancestry was associated with 15% higher odds of HTN, with corresponding increases of 0·9 mmHg and 0·7 mmHg in SBP and DBP, respectively (p < 0·001). In contrast, each 10% increase in AI ancestry was associated with 12% lower odds of HTN, along with decreases of 0·4 mmHg and 0·6 mmHg in SBP and DBP, respectively (p < 0·001). **(Table 3).** Lastly, HTN prevalence was greater (with more AA and less AI ancestry) among those reporting *More than one* race vs. those reporting race as *Unknown/Not reported/Refused* (p<0·05). **(eTable 6)**

**Table 2.** Hypertension Prevalence and Blood Pressure according to Self-reported Race and Genetic Ancestry.

| Self-Reported Race<br>(N=16,316) |  |  | Hypertension |  | Blood Pressure, mean (mmHg) |  |  |  |  |  |
| --- | --- | --- | --- | --- | --- | --- | --- | --- | --- | --- |
|  |  |  | % | p* | Systolic | C.I. | p† | Diastolic | C.I. | p† |
| White |  |  | 43.4 | <0.001 | 121.1 | 120.4-121.9 | <0.001 | 72.9 | 72.4-73.4 | <0.001 |
| Black |  |  | 45.6 | <0.001 | 124.2 | 122.3-126.2 | <0.001 | 75.3 | 74.0-76.5 | <0.001 |
| Native American |  |  | 37.6 | 0.32 | 121.5 | 119.6-123.5 | <0.05 | 72.5 | 71.2-73.8 | 0.12 |
| Multiple/Missing |  |  | 34.5 | REF | 118.7 | 118.1-119.2 | 1 | 71.5 | 71.1-71.9 | 1 |
| Overall |  |  | 38.5 |  |  |  |  |  |  |  |
| Genetic Ancestry<br>(N=10,586) |  |  | Hypertension |  | Blood Pressure, mean (mmHg) |  |  |  |  |  |
|  |  |  | % | p‡ | Systolic | C.I. | p§ | Diastolic | C.I. | p§ |
| AA | <25.4 | <80th | 39.4 | <0.001 | 120.4 | 119.8-121.1 | <0.001 | 72.2 | 71.8-72.7 | <0.001 |
|  | ≥25.4 | ≥80th | 48.0 |  | 123.2 | 122.1-124.3 |  | 74.6 | 73.8-75.4 |  |
| EA | <78.2 | <80th | 37.7 | <0.001 | 120.1 | 119.5-120.7 | <0.001 | 72.1 | 71.7-72.5 | <0.001 |
|  | ≥78.2 | ≥80th | 54.8 |  | 124.5 | 123.3 - 125.6 |  | 74.9 | 74.2 - 75.6 |  |
| AI | <51.2 | <80th | 44.7 | <0.001 | 121.9 | 121.3-122.6 | <0.001 | 73.4 | 73.0-73.8 | <0.001 |
|  | ≥51.2 | ≥80th | 26.7 |  | 117.2 | 116.3-118.2 |  | 69.8 | 69.1-70.5 |  |

P-values of \*chi2 for two proportions and †Student t-test for two means, with Unknown as reference category. C.I.: confidence interval. P-values of ‡chi2 for two proportions and §Student t-test for two means; AA: African ancestry; EA: European ancestry; AI: Amerindian ancestry. Percentages and means are weighted.

**Table 3.** Association of Percent Genetic Ancestry with Hypertension Prevalence and Blood Pressure (N=10,586)

| Genetic Ancestry | HTN prevalence |  | SBP |  | DBP |  |
| --- | --- | --- | --- | --- | --- | --- |
|  | OR* | p-value | Beta† | p-value | Beta† | p-value |
| **Crude** |  |  |  |  |  |  |
| AA | 1·11 | <0·001 | 0·9 | <0·001 | 0·7 | <0·001 |
| EA | 1·15 | <0·001 | 0·8 | <0·001 | 0·6 | <0·001 |
| AI | 0·83 | <0·001 | -1·1 | <0·001 | -0·8 | <0·001 |
| **Age and sex adjusted** |  |  |  |  |  |  |
| AA | 1·15 | <0·001 | 0·9 | <0·001 | 0·7 | <0·001 |
| EA | 1·06 | <0·001 | -0·03 | 0·69 | 0·3 | <0·001 |
| AI | 0·88 | <0·001 | -0·4 | <0·001 | -0·6 | <0·001 |

AA: African ancestry; EA: European ancestry; AI: Amerindian ancestry; HTN: hypertension; SBP: systolic blood pressure; DBP: diastolic blood pressure; OR: odds ratio; Beta: beta-coefficient of the \*logistic and †linear regression analyses of each genetic ancestry (unit in 10% change) independently.

## DISCUSSION

Given that over half of Hispanics/Latinos self-identified race as Multiple/Missing (*More than one race*, *Unknown, Not reported*, or *Refused*), our population-based study offers a sociological perspective on the utilization of genetic ancestry for HTN risk characterization. HTN prevalence and BP levels were highest among Hispanic/Latino individuals self-reporting Black race and those in the highest AA quintiles. In contrast, HTN prevalence and BP levels were lowest among individuals of Native American race and those in the highest AI quintile. Our results suggest that although the current concept of self-reported race does not fully capture the racial identity of the majority of Hispanic/Latino people, genetic ancestry may complement self-reported race in characterizing HTN risk in Hispanic/Latino individuals.

### Race as a complex social construct for Hispanics/Latinos

Nearly one-third of Hispanic/Latino individuals self-reported race as *Unknown/Not reported/Refused*, concordant with reports about the imprecision of racial self-identification among Hispanics/Latinos according to US concepts.^4^ ^14^ Hispanic/Latino racial diversity resulted from the admixture of three founding populations within a relatively short temporal window,^2^ with subsequent development of a social context of race in Latin America that differs from the US. While the US developed rigid binary racial laws^15^ motivated by economic restrictions, Latin American countries developed more fluid systems^16^ with non-legal intermediate racial categories based on societal hierarchy, not economic laws. Both contexts preserve racist ideologies, but with different consequences in Latin America vs. the US, such as the earlier abolishment of slavery, early acceptance of interracial relationships, and the lack of racial segregation laws.^17^ Despite its imperfections,^18^ self-reported race is a construct that has contributed to our understanding of health inequities in the US^19^ and should evolve to more adequately appreciate health disparities within the Hispanic/Latino population.^20,21^

### Hypertension-related environmental and genetic interplay in Hispanics/Latinos

HTN is a complex condition influenced by genetic, environmental, and lifestyle factors. Genetic ancestry patterns revealed remarkable genetic heterogeneity within US Hispanic/Latino populations,^10,22^ and may capture sociological factors particular to this group,^4,6,23^ which can impact biological outcomes. The associations between genetic ancestry and HTN found in our study are unlikely to be explained by BP-related genetic variants alone, as HTN differences across genetic ancestry groups are likely impacted by social context, such as structural racism. For example, greater African genetic ancestry was not associated with resistant HTN in a Brazilian population,^24^ but was linked to higher HTN risk in a US study.^25^ Genetic ancestry groups may overlap with self-reported race with regard to SDoH,^26^ and this intersectionality deserves future study to establish sociological factors as intermediaries between genetic ancestry and biological/health outcomes.

### Race and genetic ancestry among specific Hispanic background groups

Our analysis reiterates that Mainland Hispanics had higher AI and less AA ancestry than Caribbean Hispanics, while Cubans had more EA than other groups, consistent with reports from the US^22,27^ and Latin America.^28^ Clearly, the evolution of population genetics in Latin America is strongly tied to its anthropological colonial history.^29^ This pattern likely reflects the history of devastation of indigenous groups in the Caribbean^30^ and the forced migration of West Africans to the Caribbean through the trans-Atlantic slave trade.^31^ On the other hand, Mainland Hispanics come from regions where prominent pre-Columbian civilizations developed,^32^ with features that could have mitigated the detrimental impact of conquest on their indigenous populations.^33^ Although it is unclear, the high representation of self-reported White race and EA ancestry among Cuban heritage individuals found in our study possibly reflects differences in the colonial history of Cuba,^34^ the demographics of the migration waves of those who fled Cuba to the US,^35^ and the recruitment methods in this study.

### Black Hispanics, African ancestry predominance, and worse HTN traits

Our results show that self-reported Black race is associated with AA ancestry. Hispanics/Latinos self-reporting Black race had a majority proportion of AA ancestry (55·7%, SE ±1·4) and were highly represented (91·7%) in the top AA quintile. Self-reported Black race was associated with a worse HTN profile. Limited data suggest that while cultural and lifestyle health behaviors among Hispanic/Latino individuals self-reporting Black race may resemble those of other Hispanics/Latinos,^36^ some health metrics (e.g., self-reported health) are more similar to non-Hispanic Blacks.^37^ For example, Caribbean-Hispanics have a higher HTN prevalence^38^ compared to non-Hispanic Whites.^39^ This intersectionality of race, genetic ancestry, and health outcomes disparities deserves further study.

### White Hispanics and EA ancestry are associated with worse HTN traits

We showed an association between self-reported White race and EA ancestry in Hispanics/Latinos. Additionally, self-reported White race was associated with increased HTN prevalence and BP levels, compared to the Multiple/Missing self-reported race group with higher mean AI proportions. Prior studies have shown an association between EA and lower SBP compared with AA,^25,40^ but none have compared EA and AI.

### Amerindian Ancestry and the Hispanic/Latino Paradox

Our results suggest an association between AI ancestry with favorable BP levels and HTN prevalence, compared with AA and EA. In Brazil, higher AI ancestry was associated with increased diabetes prevalence,^41^ while in US veterans it was associated with lower DBP.^25^ In MESA, Hispanics/Latinos of Mexican heritage had increased diabetes prevalence with lower HTN prevalence compared to Caribbean Hispanics. Although genetic regions linked to AI ancestry have been identified that may explain differences in BP traits,^42,43^ further research is needed. Our results may provide insight into the Hispanic Paradox, originally reported among people of Mexican descent.^44^ This paradox, although controversial,^45^ refers to the observation of lower all-cause and cardiovascular mortality among Hispanics/Latinos persons compared to non-Hispanic Whites, despite lower SES and higher comorbidities. While speculative, Hispanic/Latino people with high AI ancestry may have some evolutionary adaptations to the Americas that could partially explain better health outcomes.^40^ Our results suggest that identifying genetic ancestry and disease-related genetic variants may provide insights into mechanisms underlying the Latino Paradox that are not reflected by self-reported race and ethnicity alone. Also, while SDoH has relatively strong associations with HTN prevalence compared to genetic determinants, limited studies exist among Hispanics/Latinos^46,47^ and further research is needed.

### Complementary Role of Genetic Ancestry and Race

Self-reported race alone provides an incomplete understanding of HTN prevalence among Hispanics/Latinos, given that one-third self-reported their race as *Unknown/Not reported/Refused*. In this context of missingness, genetic ancestry can provide additional insight, as exemplified by our finding that greater AA and AI ancestry are associated with less favorable and more favorable HTN profiles, respectively. However, previous studies on HTN disparities have relied on self-reported race, which is easier to collect in large populations than genetic ancestry analysis. Therefore, self-reported race remains useful, albeit with important limitations among Hispanic/Latino people. We show that self-reported race and genetic ancestry are complementary in providing a more complete HTN risk characterization of the Hispanic/Latino community.

### The connotation of Multiple/Missing self-reported race

Among those who reported *More than one race*, we do not have granular data as to how many or which specific races they identify with. Notably, people who self-reported their race were allowed, by design, to select only one category; no responses overlapped. Thus, the intention of those who answered *More than one race* cannot be determined with certainty. It may be that they are 1) acknowledging that they have a parent or grandparent who is of non-Hispanic heritage; 2) acknowledging that Hispanics/Latinos are a highly admixed population, with AA, EA, and/or AI lineage all within their family tree, and thus may not identify with a single race; 3) uncertain regarding where they fit within the US racial categorization scheme, for similar reason as those 32·7% who self-reported race as *Unknown/Not reported/Refused.* These reasons outline the rationale for the composite Multiple/Missing self-reported race category.

### Revised Statistical Policy Directive 15

Since 1977, the US has standardized race and ethnicity data across federal datasets^48^ and modified these categorizations accordingly as the US population has changed significantly over time. In 2024, the US revised these categorizations^48^ by combining race and ethnicity into a single question that allows for selection of Hispanic/Latino as a single merged race/ethnicity category and for selection of multiple races, aiming to reduce non-responses among Hispanic/Latino populations.^49,50^ We feel these changes inadequately capture the racial heterogeneity within Hispanics/Latinos for several reasons: 1) Race and ethnicity are two different entities, as shown in our disaggregated analysis; 2) The option *More than one race* is non-informative without additional detail, as explained in the prior section; and 3) Hispanic/Latino background group only roughly reflects the variation in self-reported race and genetic ancestry within each group. The new standards may capture the racial heterogeneity among non-Hispanic populations, since race is more consistently operationalized for these groups in the US, but they will not work for identifying people at high disease risk among Hispanic/Latino populations. A better approach than the revised standards may combine country of origin, self-reported race, ethnicity, and genetic ancestry, but this is unclear, and more research is needed. Improving the Statistical Policy Directive 15 will require ongoing monitoring and broader input from Hispanics/Latinos of diverse backgrounds, races, and genetic ancestries, particularly those who self-identify as Black race or with predominant AI genetic ancestry.

### Strengths and Limitations

Our study utilized the HCHS/SOL, the largest and most comprehensive population-based cohort of Hispanic/Latino individuals in the US, and is the first to examine how self-reported race and genetic ancestry relate to HTN risk across Hispanic/Latino background groups. However, there are limitations. 1) HCHS/SOL primarily includes urban and lower SES participants, which may limit the generalizability to the broader US Hispanic/Latino population. 2) We used linear and logistic regression with one ancestry group per model; testing ancestry interactions and collinearity were beyond the scope of this study. 3) Our measures were conducted using the 2008-2011 standardized race and ethnicity questions; future studies can consider using the new standards. 4) Black is conflated with African American in the questionnaire language; Hispanics will obviously not identify as African American; more appropriate language would have been to state “Black or Afro Latino.” 5) Future research should examine how genetic ancestry relates to social factors such as healthcare access, and how socioeconomic status, acculturation, and genetic ancestry interrelate to influence health outcomes among Hispanics/Latinos. We hope our study lays the groundwork for complementary use of self-reported race and genetic ancestry to characterize cardiovascular risk among Hispanics/Latinos and guide effective preventive strategies.

## CONCLUSION

Nearly one-third of Hispanic/Latino people did not self-report race, reinforcing the notion that US metrics imprecisely capture the social context of race in this population. Hispanics/Latinos self-reporting race as Black or White had predominantly AA or EA ancestry, respectively. Self-reported Black race and the highest AA quantile, vs. self-reported Native American race and the highest AI quantile, exhibited the least and most favorable HTN profiles, respectively, supporting the view that both genetic ancestry and the social context of race can complement our understanding of differential HTN traits in this admixed population.

## Data Availability

All data were publicly available, de-identified, and obtained in aggregate; institutional review board approval was deemed not required. Data will be available on request.

## ACKNOWLEDGEMENTS

The authors thank the HCHS/SOL staff and participants for their important contributions. Investigators website - http://www.cscc.unc.edu/hchs/

## FUNDING

The Hispanic Community Health Study/Study of Latinos is a collaborative study supported by contracts from the National Heart, Lung, and Blood Institute to the University of North Carolina (75N92025D00001, HHSN268201300001I / N01-HC-65233), University of Miami (75N92025D00009, HHSN268201300004I / N01-HC-65234), Albert Einstein College of Medicine (75N92025D00007, HHSN268201300002I / N01-HC-65235), University of Illinois at Chicago (75N92025D00008, HHSN268201300003I / N01-HC-65236 Northwestern Univ), and San Diego State University (75N92025D00010, HHSN268201300005I / N01-HC-65237). The following Institutes/Centers/Offices have contributed to the HCHS/SOL through a transfer of funds to the NHLBI: National Institute on Minority Health and Health Disparities, National Institute on Deafness and Other Communication Disorders, National Institute of Dental and Craniofacial Research, National Institute of Diabetes and Digestive and Kidney Diseases, National Institute of Neurological Disorders and Stroke, NIH Institution-Office of Dietary Supplements.

## DISCLOSURES

First and senior authors had full access to the study data and were responsible for data integrity and the accuracy of the analyses. All authors have reviewed and approved the final manuscript. None of the authors had any financial or other conflicts of interest.

## SUPPLEMENTARY MATERIAL

**eTable 1.** Baseline Characteristics of HCHS/SOL Study Population according to Self-Reported Race.

**eTable 2.** Background group among Multiple/Missing Self-reported Race, per Sub-Components.

**eTable 3.** Proportion of Genetic Ancestry Quintiles within Self-Reported Race.

**eTable 4.** Genetic Ancestry per Background Group and Self-Reported Race (N=10,586).

**eTable 5.** Genetic Ancestry and Hypertension (AHA/ACC), per several ancestry thresholds.

**eTable 6.** Genetic Ancestry among Multiple/Missing Self-reported Race, per Sub-Component.

## Notes

### Competing Interest Statement

The authors have declared no competing interest.

### Author Declarations

All data were publicly available, de-identified, and obtained in aggregate; institutional review board approval was deemed not required.

